# Shared genetic etiology and causality between COVID-19 and venous thromboembolism: evidence from genome-wide cross trait analysis and bi-directional Mendelian randomization study

**DOI:** 10.1101/2022.05.21.22275413

**Authors:** Xin Huang, Minhao Yao, Peixin Tian, Jason Y.Y. Wong, Zilin Li, Zhonghua Liu, Jie V. Zhao

## Abstract

Venous thromboembolism (VTE) occurs in up to one third patients with COVID-19. VTE and COVID-19 may share a common genetic architecture, which has not been clarified yet. To fill this gap, we leveraged summary-level genetic data from the latest COVID-19 host genetics consortium and UK Biobank and examined the shared genetic etiology and causal relationship between COVID-19 and VTE. The cross-trait analysis identified 8, 11, and 7 shared loci between VTE and severe COVID-19, COVID-19 hospitalization, SARS-CoV-2 infection respectively, in 13 genes involved in coagulation and immune function and enriched in the lung. Co-localization analysis identified eight shared loci in *ABO, ADAMTS13* and *FUT2* genes. Bi-direction Mendelian randomization suggested that VTE was associated with higher risks of all COVID-19 related traits, and SARS-CoV-2 infection was associated with higher risk of VTE. Our study provided timely evidence and novel insights into the genetic etiology between COVID-19 and VTE.

## Introduction

Coronavirus disease 2019 (COVID-19), caused by the severe acute respiratory syndrome coronavirus 2 (SARS-CoV-2) infection, has led to a worldwide pandemic since March 2020, and caused repeated waves of outbreaks across the globe. Venous thromboembolism (VTE) is one common and serious comorbidity. Notably, thrombotic events occur in up to one-third of patients with COVID-19.^1^ People with more pronounced thrombotic symptoms are more likely to develop severe COVID-19,^2^ and thrombotic complications are a well-established predictor of mortality in people with COVID-19.^1^ Consistently, several genome-wide association studies (GWASs) of COVID-19 related traits (severe COVID-19, COVID-19 hospitalization and SARS-CoV-2 infection) have identified genetic loci in *ABO*,^3-7^ an established gene related to thrombosis. These evidence suggests that VTE and COVID-19 may share common genetic architecture. Identifying shared genetic factors contributing to both COVID-19 and VTE can provide novel insights into disease pathogenesis and pinpoint targets for therapeutic development or drug repurposing. However, to our knowledge, this problem has not been comprehensively investigated.

Meanwhile, the causal relationship between VTE and COVID-19 has not been clarified. On the one hand, VTE predicts COVID-19 severity and mortality.^1 2^ On the other hand, the cytokine storm and excessive inflammation caused by SARS-CoV-2 infection is hypothesized to lead to the systemic coagulation dysfunction;^8-11^ therefore, SARS-CoV-2 infection may increase the risk of VTE. Many institutional evidence-based guidelines supported the use of prophylactic treatments such as anticoagulants for thromboprophylaxis in COVID-19 patients.^12^ However, the recommendation is mainly based on observational studies, which might be subject to unmeasured confounding bias. To resolve this issue, Mendelian randomization (MR) study uses genetic variants as instrument for causal inference and can provide unbiased causal effect estimate even in the presence of unmeasured confounding.^13^ One previous MR study in a relatively smaller GWAS for COVID-19 suggested that genetically predicted VTE is associated with higher risk of COVID-19 hospitalization and SARS-CoV-2 infection,^14^ however, the association with severe COVID-19 is uncertain possibly due to the limited sample size. The reverse association, i.e., the association of COVID-19 with VTE has not been examined. The recently updated GWAS for COVID-19 with doubled sample size provided a well-powered study dataset to investigate the relationship between VTE and COVID-19.

Taken together, to fill this knowledge gap, we performed a genome-wide cross-trait analysis for VTE and three COVID-19 related traits to estimate their genetic correlations and shared genetic components using publicly available summary level data from GWASs of VTE and COVID-19. We also applied bi-directional Mendelian randomization methods to study the causal relationship between VTE and three COVID-19 related traits.

## Results

### Genetic correlation of VTE with COVID-19 related traits

The heritability (h^2^) estimated by Linkage disequilibrium score regression (LDSC) analysis suggested severe COVID-19, COVID-19 hospitalization, SARS-CoV-2 infection and VTE are heritable (P<0.05 shown in Supplementary Table 1). We found positive overall genetic correlation of VTE with COVID-19 hospitalization (rg = 0.2320, P-value= 0.0092) (shown in Figure 1 and Supplementary Table 2). The genetic correlation of VTE and severe COVID-19 and SARS-CoV-2 were positive but not significant at p value of 0.05 level. Further partitioned LDSC analysis found that severe COVID-19, COVID-19 hospitalization and SARS-CoV-2 infection are genetically correlated with VTE in 5 (Fetal DNase I hypersensitivity sites (DHS), H3K4me1, H3K9ac H3K27ac and transcription factor-binding site (TFBS)) (Figure 2), 11 (except for super enhancers regions) and 10 (except for conserved and super enhancers regions) of the 12 functional categories, respectively.

**Figure 1.**
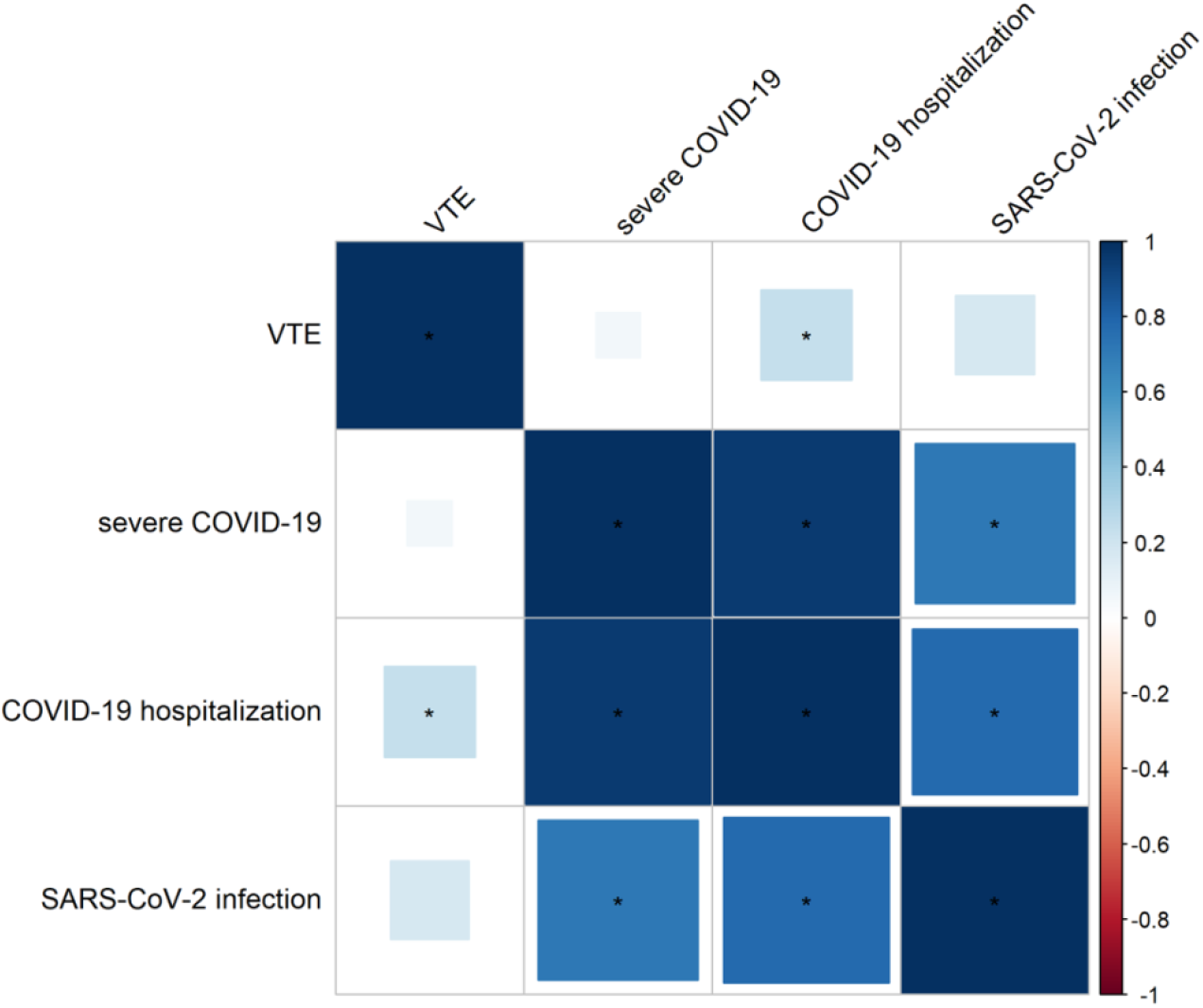
Genetic correlation of VTE with COVID-19 related traits.

**Figure 2.**
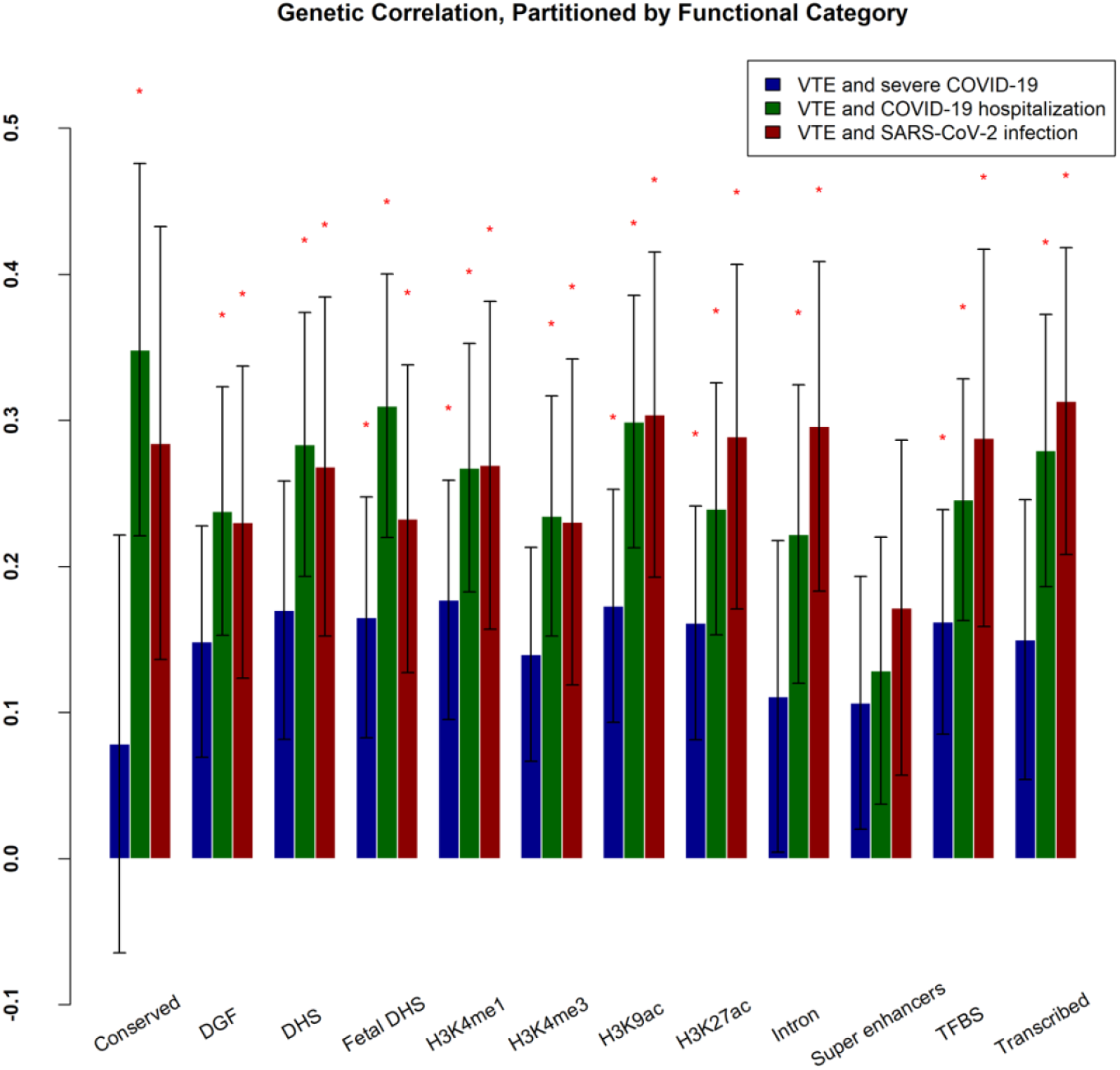
Partitioned genetic correlation between VTE and COVID-19 related traits. The x-axis represents the 12 functional categories and the y-axis represents the estimated partitioned genetic correlation. The significant functional categories (P<0.05) are starred. DGF: DNaseI digital genomic footprinting; DHS: DNase I hypersensitivity site; TFBS: transcription factor binding sites.

### Multi-Trait Analysis of GWAS (MTAG)

Based on the COVID-19 host genetics initiative (HGI) updated GWAS meta-analysis (release 7), there were 45 genome-wide significant (P<5×10^−8^) and uncorrelated (r^2^<0.01) loci for severe COVID-19, 46 for COVID-19 hospitalization and 26 for SARS-CoV-2 infection (Supplementary Table 3), and 12 for VTE in the GWAS from UK Biobank (Supplementary Table 4). Compared with these previously identified SNPs, using MTAG incorporating information from GWAS of COVID-19 and VTE we did not identify additional genome-wide significant loci for COVID-19 traits or VTE.

As shown in Table 1, we identified 8 shared novel genetic loci associated with both VTE and severe COVID-19, 11 with both VTE and COVID-19 hospitalization and 7 with both VTE and SARS-CoV-2 infection (Pmeta < 5×10^−8^; single trait P <0.05). Figure 3 displays the Manhattan plots of these results. In line with previous studies,^3-6,15,16^ we identified *ABO* gene and *FUT2* gene, which contributed to both VTE and COVID-19. Notably, we identified seven novel genes which have not been reported yet, including *LINC00970*, and six protein-coding genes (*TSPAN15, ADAMTS13, F5, DNAJB4, SLC39A8* and *OBSCN)*.

**Table 1.** Genome-wide significant loci associated with the COVID-19 traits and VTE in cross trait meta-analysis.

| SNP | CHR | A1 | A2 | Representative gene | P_1 | P_2 | P_META | Coloc H4 Prob |
| --- | --- | --- | --- | --- | --- | --- | --- | --- |
| Severe COVID-19 and VTE |  |  |  |  |  |  |  |  |
| rs114101204 | 1 | A | G | LINC00970 | 2.16E-47 | 3.39E-02 | 1.44E-37 | 2.25E-04 |
| rs7726201 | 5 | T | C | nan | 2.28E-02 | 1.23E-07 | 2.17E-08 | 2.00E-02 |
| rs11244061 | 9 | T | C | ABO | 2.66E-26 | 1.81E-04 | 3.55E-26 | 8.55E-01 |
| rs149181677 | 9 | T | C | ADAMTS13 | 1.39E-08 | 7.36E-04 | 2.07E-10 | 8.55E-01 |
| rs138683771 | 9 | C | A | ABO | 6.77E-12 | 7.22E-03 | 1.42E-09 | 8.55E-01 |
| rs75583483 | 10 | G | A | TSPAN15 | 1.73E-15 | 4.23E-02 | 6.04E-14 | 3.19E-02 |
| rs676314 | 19 | G | A | NAPSA | 4.44E-02 | 1.42E-09 | 8.13E-10 | 1.54E-02 |
| rs2834166 | 21 | A | C | IFNAR2 | 3.25E-02 | 4.74E-14 | 4.12E-13 | 2.33E-03 |
| COVID-19 hospitalization and VTE |  |  |  |  |  |  |  |  |
| rs114101204 | 1 | A | G | LINC00970 | 2.16E-47 | 9.26E-03 | 1.65E-25 | 1.29E-03 |
| rs9287090 | 1 | A | G | F5 | 7.69E-18 | 2.93E-02 | 1.61E-11 | 1.98E-03 |
| rs34517439 | 1 | A | C | DNAJB4 | 4.50E-02 | 3.41E-09 | 4.96E-09 | 5.94E-03 |
| rs7532924 | 1 | A | C | OBSCN | 2.84E-02 | 2.76E-07 | 2.97E-08 | 8.91E-02 |
| rs13135092 | 4 | G | A | SLC39A8 | 2.04E-02 | 1.49E-07 | 1.42E-08 | 4.94E-02 |
| rs11244061 | 9 | T | C | ABO | 2.66E-26 | 8.70E-10 | 3.64E-28 | 9.00E-01 |
| rs8176686 | 9 | C | T | ABO | 1.40E-09 | 1.63E-07 | 1.89E-14 | 9.00E-01 |
| rs149181677 | 9 | T | C | ADAMTS13 | 1.39E-08 | 7.13E-05 | 1.09E-10 | 9.00E-01 |
| rs199501 | 17 | G | A | WNT3 | 3.52E-02 | 1.45E-13 | 6.84E-10 | 8.26E-03 |
| rs676314 | 19 | G | A | NAPSA | 4.44E-02 | 8.95E-15 | 4.10E-15 | 1.55E-02 |
| rs2248202 | 21 | A | C | IFNAR2 | 3.45E-02 | 4.45E-15 | 3.04E-15 | 2.32E-03 |
| SARS-CoV-2 infection and VTE |  |  |  |  |  |  |  |  |
| rs76896797 | 3 | T | C | SLC6A20 | 3.42E-02 | 4.63E-15 | 2.15E-11 | 3.60E-03 |
| rs13107325 | 4 | T | C | SLC39A8 | 2.69E-02 | 5.14E-08 | 9.54E-09 | 4.81E-02 |
| rs550057 | 9 | T | C | ABO | 4.25E-38 | 4.05E-78 | 1.56E-51 | 9.84E-01 |
| rs9411367 | 9 | T | C | ABO | 4.64E-05 | 8.09E-11 | 3.20E-12 | 9.84E-01 |
| rs71503180 | 9 | A | G | ABO | 2.07E-04 | 6.81E-08 | 3.05E-10 | 9.84E-01 |
| rs492602 | 19 | G | A | FUT2 | 9.18E-06 | 7.26E-15 | 1.42E-10 | 8.18E-01 |
| rs676314 | 19 | G | A | NAPSA | 4.44E-02 | 1.02E-08 | 3.54E-09 | 1.54E-02 |

CHR, chromosome; A1, other allele; A2, effect allele
P\_1 represent the p-values in the GWAS of VTE. P\_2 represent the p-values in the GWASs of COVID-19 related traits. P\_META represents the p-value of the cross-trait meta-analysis.

**Figure 3.**
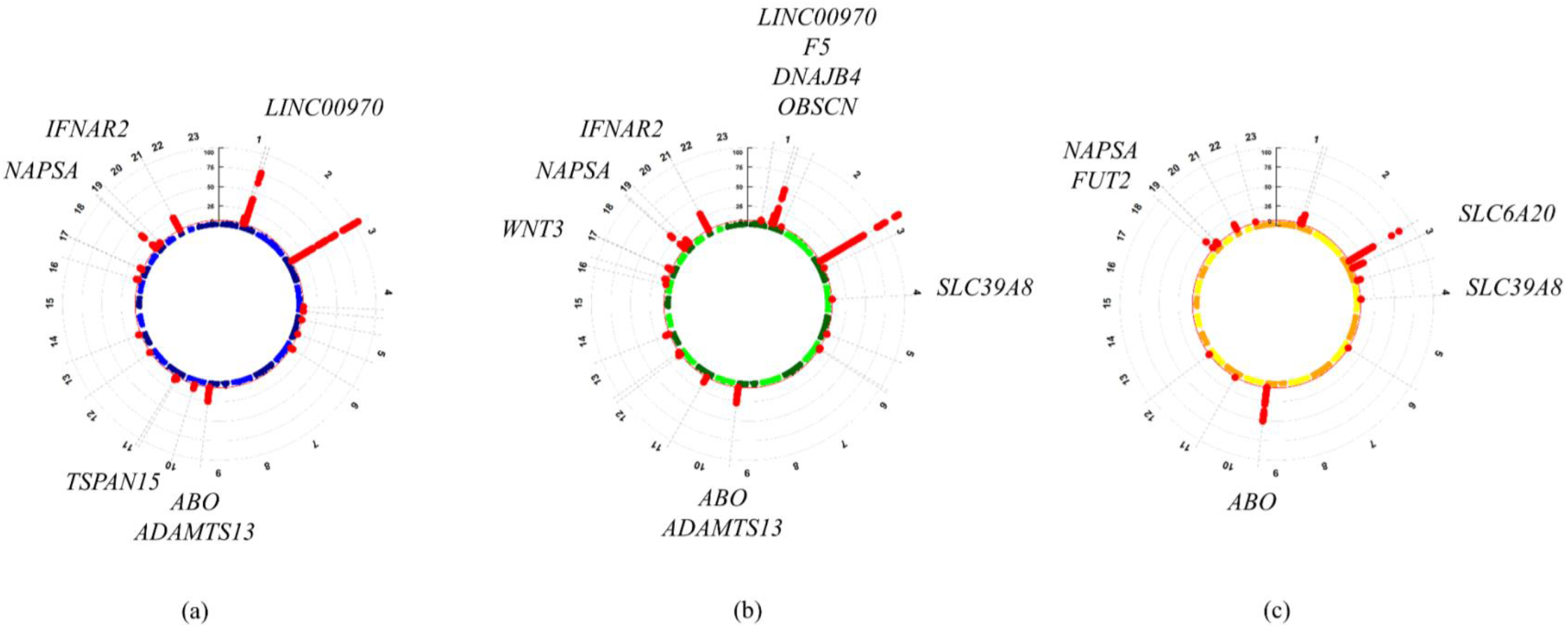
Circular Manhattan plot of cross-trait analysis for (a) VTE and severe COVID-19, (b) VTE and COVID-19 hospitalization, (c) VTE and SARS-CoV-2 infection. Each point represents a SNP, and shared significant loci with meta-analysis P-value < 5×10^−8^ and single-trait P-value < 0.05 are colored in red. SNPs are arranged according to the chromosome position.

For severe COVID-19 with VTE, the strongest association signals were localized to the *LINC00970* gene (index SNP: rs114101204 for severe COVID-19 and VTE, Pmeta=1.44×10^−37^) at locus 1q24.2. For COVID-19 hospitalization and SARS-CoV-2 infection with VTE, the strongest association signals were localized on or near the *ABO* gene (index SNP: rs11244061 for COVID-19 hospitalization, Pmeta=3.64×10^−28^; rs550057 for SARS-CoV-2 infection, Pmeta=1.56×10^−51^) at locus 9q34.2.

### Fine-mapping and colocalization analysis identify shared causal variants

For each of the shared loci of VTE with COVID-19, Supplementary Tables 5-7 listed all SNPs within 500 kb of these variants in the 99% credible sets. Co-localization analysis shows that those colocalized genetic loci for VTE and severe COVID-19 were located in *ABO* (index SNPs: rs11244061 and rs138683771), *ADAMTS13* (index SNP: rs149181677), the colocalized loci for VTE and COVID-19 hospitalization were located in *ABO* (index SNPs: rs11244061and rs8176686) and *ADAMTS13* (index SNP: rs149181677), the colocalized loci for VTE and SARS-CoV-2 infection were located in *ABO* (index SNPs: rs550057, rs9411367 and rs71503180) and *FUT2* (index SNP: rs492602) (Table 1).

### GTEx tissue-specific expression analysis (TSEA) and over-representation enrichment analysis of shared genes

The identified shared genes for severe COVID-19 and COVID-19 hospitalization with VTE were significantly enriched for expression in the lung tissue; however, no highly enriched tissues were found for SARS-CoV-2 infection and VTE (Figure 4). Gene ontology (GO) analysis highlighted several significant shared biological processes between three COVID-19 traits and VTE, such as “calcium-mediated signaling”, “second-messenger-mediated signaling”, “chemokine-mediated signaling pathway”, “response to chemokine”, “response to type I interferon” (Supplementary Tables 8-10).

**Figure 4.**
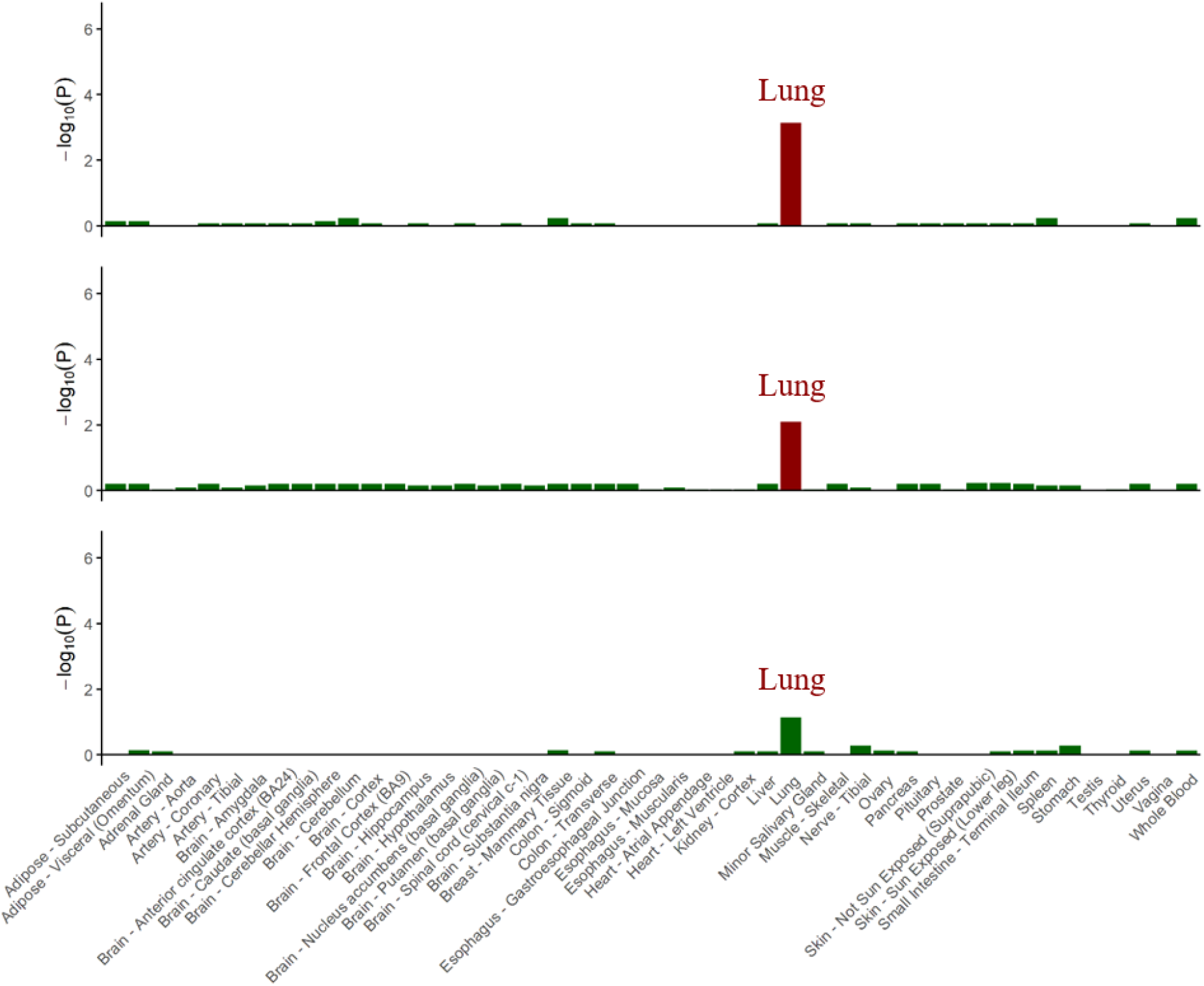
GTEx tissue enrichment analysis for expression of shared significant genes (meta-analysis P-value < 5×10^−6^) for (a) VTE and severe COVID-19, (b) VTE and COVID-19 hospitalization, (c) VTE and SARS-CoV-2 infection. P-values of Fisher’s exact test after Benjamin-Hochberg correction are presented in – log10 scale. Red represents significant tissue enrichment (Lung, P-value= 7.16×10^−4^ for severe COVID-19 and VTE, P-value= 8.20×10^−3^ for COVID-19 hospitalization and VTE).

### Bi-directional MR analysis

We found that genetically predicted VTE was positively associated with higher risk of all three COVID-19 related traits (OR= 1.08 for severe COVID-19, 95% CI: 1.04-1.12, P-value= 3.22×10^−5^; OR=1.08 for COVID-19 hospitalization, 95% CI: 1.05-1.11, P-value =9.09×10^−10^; OR=1.05 for SARS-CoV-2 infection, 95% CI: 1.04-1.07, P-value= 1.02×10^−9^) (Figure 5). In the reverse direction, we found that a positive association of genetically predicted SARS-CoV-2 infection with higher risk of VTE (OR=1.80, 95% CI: 1.41-2.29, P-value=1.98×10^−6^), and null associations of genetically predicted severe COVID-19 or COVID-19 hospitalization with VTE (Figure 6). These results are robust to different MR methods (Supplementary Figures 1 and 2).

**Figure 5.**
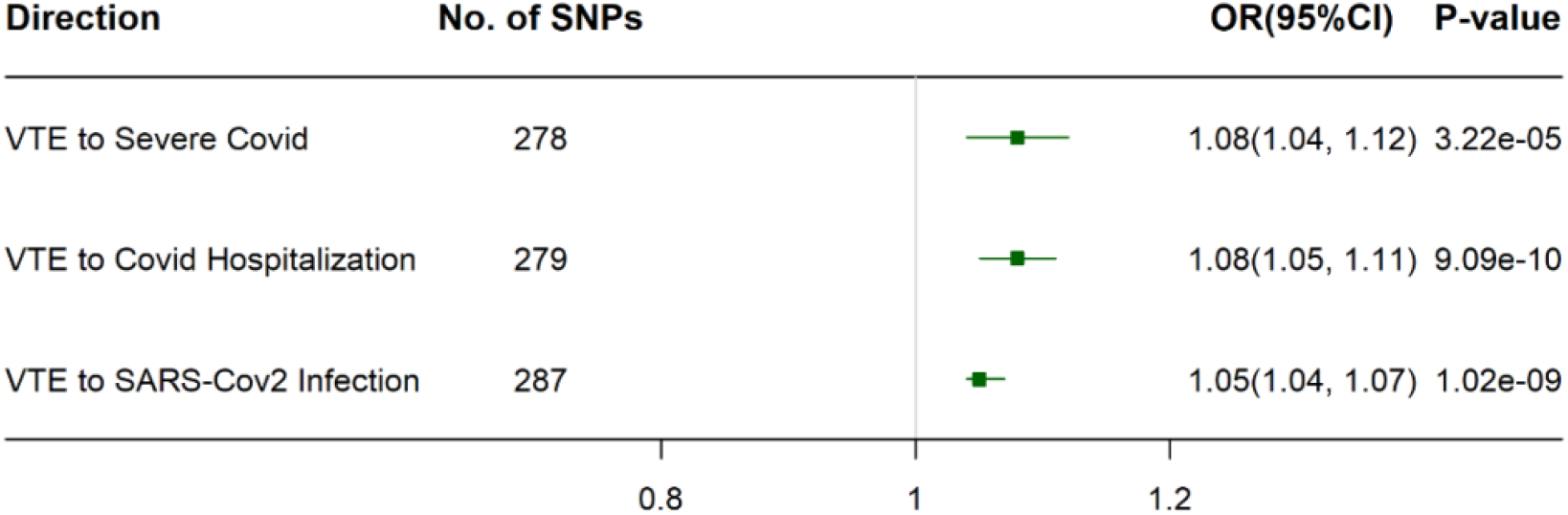
Mendelian randomization analysis on the association of genetically predicted VTE with the risk of COVID-19 related traits using inverse variance weighting (IVW). The estimates are presented as odds ratios (OR) with 95% confidence intervals (CI).

**Figure 6.**
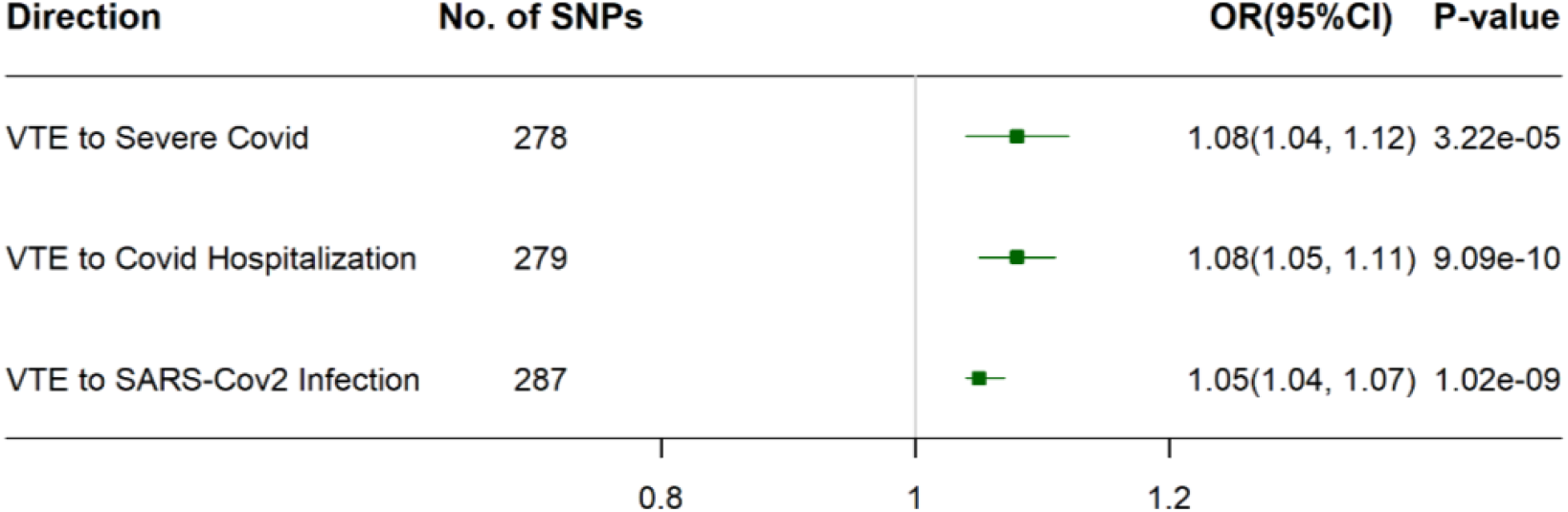
Mendelian randomization analysis on the association of genetically predicted COVID-19 related traits with the risk of VTE using IVW. The estimates are presented as odds ratios (OR) with 95% confidence intervals (CI). An arrow represents the estimate is out of boundary in this direction.

## Discussion

In this study, we comprehensively investigated the shared genetic etiology between three COVID-19 related traits and VTE based on the latest data from COVID-19 HGI and UK Biobank. For the first time, we identified 8 shared loci for severe COVID-19, 11 loci for COVID-19 hospitalization and 7 loci for SARS-CoV-2 infection with VTE, with mapped genes involved in coagulation and immune function. Co-localization analysis identified eight shared genetic loci in *ABO, ADAMTS13* and *FUT2* genes. Enrichment analysis suggested that those gene expressions were enriched mainly in the lung tissue, and supported pathways related to coagulation and immunity. Moreover, we examined their causal relationship using bi-directional MR, which suggested that VTE may increase the risk of severe COVID-19, COVID-19 hospitalization and SARS-CoV-2 infection, and interestingly, SARS-CoV-2 infection may increase the risk of VTE.

Our study found genetic correlations between VTE and COVID-19. LDSC analysis showed a significant positive genetic correlation between VTE and COVID-19 hospitalization. Although LDSC analysis did not find significant overall genetic correlation between severe COVID-19 and VTE, however partitioned LDSC analysis found severe COVID-19 and VTE were positively correlated in some functional categories, including TFBS, Fetal DHS, H3K4me1, H3K9ac and H3K27ac, which are associated with the control of transcription and the status of *cis*-regulatory elements such as promoters and enhancers within gene regulatory regions.^17-20^

We identified shared genetic loci between VTE and COVID-19 using cross-trait meta-analysis and colocalization analysis. We identified shared loci in 13 genes (*LINC00970, ABO, ADAMTS13, TSPAN15, NAPSA, IFNAR2, F5, WNT3, DNAJB4, SLC39A8, OBSCN, SLC6A20, FUT2*) and co-localized genetic loci in three genes (*ABO, ADAMTS13* and *FUT2*), involved in coagulation and immune function. Consistent with previous report, we confirmed the role of *ABO, NAPSA, IFNAR2* in severe COVID-19 and COVID-19 hospitalization,^3,4,7,21,22^ *WNT3* in COVID-19 hospitalization^23^, and *SLC6A20* in SARS-CoV-2 infection^4^. Notably, we found several novel genes, including *ADAMTS13, TSPAN15, WNT3, DNAJB4, SLC39A8, OBSCN*, which have not been reported yet. These novel findings provide new insights into genetic etiology of both VTE and COVID-19 related traits, and may help pinpoint therapeutic targets.

Interestingly, we found several genes related to coagulation. In cross-trait meta-analysis, we found a strong shared genetic signal for severe COVID-19 and VTE located at locus 1q24.2 in *LINC00970* close to *NME7* gene. Locus 1q24.2 has been found to be associated with VTE previously,^24^ but has not been linked to severe COVID-19. This locus (1q24.2) also includes shared genetic variant in *F5* gene. *F5* gene encodes plasma procoagulant factor (F)V, a coagulation factor involved in thrombin activation, and its impaired downregulation is key to thrombosis.^25^ Similarly, *ABO, FUT2*, and *ADAMST13* are well-established VTE related genes,^24,26^ and were found in the colocalization analysis. Our findings are consistent with previous genetic studies showing *ABO* gene is associated with severe COVID-19 and SARS-CoV-2 infection,^3-5,15^ possibly by regulating thrombosis.^3,27^ In line with the role of *ABO* gene, we found that *FUT2* gene, a fucosyltransferase gene involved in ABO blood group antigen synthesis, was associated with both VTE and SARS-CoV-2 infection. We also found a novel shared gene, *ADAMST13*, which encodes protein ADAMST13. The function of ADAMTS13 might be affected by ABO blood group,^28^ and a previous MR study showed genetically predicted ADAMST13 is associated with severe COVID-19.^29^

Moreover, several shared genes are involved in immune function. The signals shared by severe COVID-19 and COVID-19 hospitalization with VTE were also mapped to the *IFNAR2* gene at 21q22.11 and *NAPSA* gene at 4q24. *IFNAR2* gene encodes interferon receptor subunit that mediates the early host immune response to viral infection.^7^ *NAPSA* gene is associated with damage-associated transient progenitors promoted by inflammation.^21,30^ Other shared genes have been known to be associated with innate antiviral defenses (*SLC6A20*),^3,31^ immune cell signaling regulation (*WNT3, TSPAN15)*^32,33^, inflammatory lung injury *(SLC39A8)*^34^ or tumor activation (*TSPAN15, DNAJB4, OBSCN*).^35-37^ These results imply that immune function may be related to both COVID-19 and VTE.

The findings from enrichment analysis also suggest immune function might be involved in the shared etiology. In the enrichment analysis, we found several shared pathways related to immune function, such as calcium-mediated signaling and chemokine/interferon related response. Previous studies suggested that calcium signaling may play a role in hemostasis and thrombosis.^38,39^ Calcium signaling is also of paramount importance in immune cells,^40,41^ and chemokine and interferon play a major role in activating host immune and inflammatory responses.^42-44^ Our TSEA reported that shared genes for severe COVID-19 and COVID-19 hospitalization with VTE were mainly enriched for gene expressions in the lung tissue. Consistently, pulmonary vascular endothelial injury and immunothrombosis (most of which occur within the lung microvessels) are key drivers of severe events after SARS-CoV-2 infection, such as acute respiratory failure and ARDS.^45,46^ So, these evidence suggests that the common pathways between COVID-19 and VTE may relate to immunity, endothelial cell function, and coagulation.

Our study also provided timely evidence regarding the causal relationship between VTE and COVID-19 traits. Interestingly, our MR analysis showed a positive association between genetic susceptibility for VTE and the risk of severe COVID-19, COVID-19 hospitalization and SARS-CoV-2 infection, as well as a positive association of SARS-CoV-2 infection with VTE. Despite the statistical significance, our finding still needs to be interpreted with caution as we cannot exclude that this positive association may have occurred by chance. However, the positive association was also shown in a previous MR study using COVID-19 HGI release 5 data showing that genetically predicted VTE was associated with higher risk of COVID-19 hospitalization and SARS-CoV-2 infection.^14^ Using COVID-19 HGI release 7 with doubled sample size of release 5, we added by showing a positive association of genetically predicted VTE with higher risk of severe COVID-19.

Our study delivers an important information that VTE possibly increase the risk of COVID-19, which matters in the consideration of COVID-19 management strategies, especially for people with VTE. For example, taking vaccination to lower the risk for severe COVID-19 in people with VTE. Meanwhile, our MR analysis suggested that genetically predicted SARS-CoV-2 infection was associated with a higher risk of VTE. Partly consistent with our results, in a cohort of 153,760 individuals with COVID-19, as well as two sets of control cohorts with 5,637,647 (contemporary controls) and 5,859,411 (historical controls) individuals, people with SARS-CoV-2 infection had higher risk of cardiovascular events, including VTE.^47^ Nevertheless, a replication study of MR analysis in a larger GWAS of VTE will be worthwhile.

Our study is the first to use large-scale genetic data to explore the shared genetic architecture between COVID-19 related traits and VTE, providing timely evidence and novel insights into the genetic etiology between them. The GWAS summary-level data for COVID-19 was obtained from the COVID19 HGI release 7 summary statistics, the largest and latest GWAS of COVID-19 available, which doubles the sample size from the previous version, and improves study power.

We also acknowledge several limitations of our study. First, the GWAS data for COVID-19 and VTE used in this study were derived from the European population, so the associations may not be generalizable to other ancestries. Second, although GWAS summary statistics conducted study-specific quality control, misclassification of COVID-19 might exist. Third, the summary statistics limit us to assess sex and age-specific genetic effects. Fourth, the GWAS of COVID-19 related traits might be conducted at different time periods, so there might be differences in the SARS-CoV-2 infection strain. However, our study does not aim to assess the association with specific strain of SARS-CoV-2 infection. Finally, although our study provides evidence of genetic correlation and genetic overlap between COVID-19 and VTE, the underlying biological mechanisms are still unclear, and further studies are still needed for validation.

In conclusion, our findings provided novel evidence of genetic correlations between severe COVID-19, COVID-19 hospitalization, SARS-CoV-2 infection and VTE, and highlighted their common genetic architecture, with shared genes closely related to coagulation and immunity. Our work contributes to the understanding of COVID-19 and VTE etiology and provides new insights into the prevention and comorbidity management of COVID-19.

## Methods

### Study population

The GWAS summary statistics for COVID-19 of European ancestry were provided by the COVID-19 host genetics initiative round 7 (https://www.covid19hg.org/results/, release date: April 08, 2022). We included three COVID-19 related traits: (1) Severe COVID-19, defined as COVID-19-confirmed individuals with very severe respiratory symptoms or those who died from the disease (up to 13,769 cases and 1,072,442 controls); (2) COVID-19 hospitalization defined as individuals who were hospitalized for related infection symptoms, with laboratory-confirmed SARS-CoV-2 infection (up to 32,519 cases and 2,062,805 controls); (3) SARS-CoV-2 infection defined as all individuals who reported positive (laboratory diagnosis, physician diagnosis or self-report) for SARS-CoV-2 infection (up to 122,616 cases and 2,475,240 controls).

For VTE, we used summary statistics on the GWAS of VTE (3,900 cases and 369,592 controls of European ancestry) from the UK Biobank. Participating cohorts of COVID-19 and VTE were described in detail in the Supplementary Note.

### Linkage disequilibrium score regression (LDSC) analysis

LDSC analysis was conducted to assess the heritability for a single trait and genetic correlations between two traits. The analysis was conducted using the LDSC software based on the GWAS summary statistics. LD scores of 1000G European ancestry was used as reference.^48^ An estimate of the heritability or genetic correlation can be obtained by regressing the χ^2^ statistics or the products of z-scores on LD scores, respectively. LDSC can also correct for the inflation of test statistics caused by polygeneicity.^49^

### Partitioned LDSC analysis

We performed partitioned LDSC analysis to estimate the genetic correlation between two traits within each of the following 12 functional categories^18^: conserved region, DNaseI digital genomic footprinting region (DGF), DNase I hypersensitivity sites (DHS), fetal DHS, H3K4me1, H3K4me3, H3K9ac and H3K27ac, intron region, super enhancers, transcription factor-binding site (TFBS) and transcribed region. The re-calculated LD scores of the SNPs classified in each specific annotation category allowed us to find out which functional categories accounted for the majority of the overall genetic correlation.

### Multi-Trait Analysis of GWAS (MTAG)

We applied MTAG to identify novel loci with strong signals for COVID-19, and to detect shared genetic variants for VTE and three COVID-19 traits.^50-52^ MTAG can improve the effect estimates for each COVID-19 trait by incorporating the weighted sum of GWAS estimates for VTE. We also used MTAG to conduct cross-trait meta-analysis, which utilizes sample size-weighted, fixed-effect model together with genetic covariance modeling from all sources to combine evidence of the association between individual variants for VTE and COVID-19.

### Fine-mapping and co-localization analysis

For the significant shared loci between two traits, we used the Bayesian fine-mapping algorithm to identify a 99%-credible set of causal variants for each of the shared loci within 500kb of the index SNP.^53^ Then, we conducted the co-localization analysis to check whether COVID-19 and VTE association signals co-localized at shared loci, by calculating the probability that two traits shared the same causal variant (P(H_4_)).^54^ If P(H_4_) is greater than 0.5 for any SNP, we labelled it as a co-localized genetic variant.

### Tissue-specific expression analysis (TSEA) and over-representation enrichment analysis

In order to test whether the shared genes are overly expressed in a specific tissue, we performed TSEA with the HUGO Gene Nomenclature Committee (HGNC) name of genes that correspond with the shared loci in cross-trait meta-analysis. We used the R package ‘deTS’, which uses GTEx RNA-seq data and the ENCODE panel as a reference panel, and calculates the corresponding z-score for each tissue.^55^ We also used the WEB-based GEne SeT AnaLysis Toolkit to further assess the overrepresented enrichment of the same shared genes in Gene ontology (GO) biological process.^56^

### Mendelian randomization (MR) analysis

We conducted a bidirectional MR analysis to assess the causal association between each COVID-19 related trait and VTE. Genetic instruments for VTE were obtained from a published genetic instruments published in a large GWAS of VTE ((23,151 cases, 553,439 controls).^57^ The GWAS provided 297 SNPs (P-value<1×10^−5^, r^2^<0.2) as instrument for VTE (Supplementary Table 11).^57^ The genetic associations of these SNPs with COVID-19 related traits were obtained from COVID-19 HGI release 7 GWAS meta-analysis, as shown in study population. Using the same process of identifying instruments, based on COVID-19 HGI release 7 GWAS meta-analysis we obtained 221, 282 and 177 SNPs as instruments for severe COVID-19, COVID-19 hospitalization and SARS-CoV-2 infection respectively (Supplementary Tables 12-14). The genetic associations of these SNPs with VTE were obtained from the GWAS of VTE (3,900 cases and 369,592 controls of European ancestry) in the UK Biobank. We used inverse variance weighted (IVW) analysis in the main analysis,^58^ and used several other MR methods that are robust to pleiotropy, including the weighted median,^59^ MR-Egger,^60^ MR-PRESSO,^61^ and MR-RAPS,^62^ in sensitivity analysis. The details of methods were provided in the Supplemental Note.

## Data Availability

Genetic associations with VTE were from the UK biobank GWAS results provided by Lee Lab (https://www.leelabsg.org/resources). Genetic associations with severe COVID-19, COVID-19 hospitalization and SARS-CoV-2 infection was obtained from COVID-19 host genetics consortium GWAS meta-analyses round 7, downloaded from https://www.covid19hg.org/results/r7/.

https://www.covid19hg.org/results/

https://www.leelabsg.org/resources

## Statements and Declarations

### Author Contributions

The manuscript was drafted by XH, with great help of JVZ and ZL (edited by JYYW and ZLL). Data analysis was conducted by MY and PT. XH and MY interpreted the results, with the help of JYYW, ZLL, ZL and JVZ. ZL and JVZ developed the study conception, designed the analysis, and critically revised the manuscript for important intellectual content. All authors read and approved the final manuscript.

### Ethics approval

The study is an analysis using publicly available summary data that does not require ethical approval.

## Acknowledgments

This research was conducted using the summary statistics from COVID-19 host genetics consortium and UK Biobank. The authors would like to thank all participants in the study and investigators for sharing the valuable data.

## Data availability

Genetic associations with VTE were from the UK biobank GWAS results provided by Lee Lab (https://www.leelabsg.org/resources). Genetic associations with severe COVID-19, COVID-19 hospitalization and SARS-CoV-2 infection were obtained from COVID-19 host genetics consortium GWAS meta-analyses round 7, downloaded from https://www.covid19hg.org/results/r7/.

